# Molecular surveillance of Falciparum malaria in Rwanda: Shifts in parasite prevalence and risk factors between the 2014–15 and 2019–20 Rwanda Demographics and Health Surveys

**DOI:** 10.64898/2026.04.01.26349976

**Authors:** Jenna Zuromski, Neeva Wernsman Young, Pierre Gashema, Vincent Iradukunda, Ntwari Jean Bosco, Jacob M Sadler, Claudia Gaither, Tharcisse Munyaneza, Sean Vincent Connelly, Lauren Lee, Varun Goel, Claude Mambo Muvunyi, Jean De Dieu Butera, Jean-Baptiste Mazarati, Jonathan J. Juliano, Jeffrey A. Bailey

## Abstract

Rwanda is a malaria endemic country and a focal point for emerging *Plasmodium falciparum* artemisinin partial resistance (ART-R). While Demographic and Health Surveys (DHS) provide both national and province-level representative data, malaria testing in Rwandan DHS (RDHS) studies has been limited to a subset of adult women and children under 5 years using RDT and/or microscopy. Recent work using ultra-sensitive quantitative real time PCR on residual dried blood spots (DBS) from the 2014-15 RDHS revealed a significantly higher P. falciparum prevalence than detected by standard DHS diagnostics. Building on this study, we analyzed 7,127 adult DBS samples collected for HIV testing in the 2019-20 RDHS to generate updated prevalence measures. We found a national *P. falciparum* infection prevalence of 7.7% (95%CI [6.8%, 8.7%]), with predominantly low-density infections (median parasitemia: 7.3 parasites/uL). We assessed covariates of *P. falciparum* malaria infection, identifying male sex, lower household wealth, lower educational achievement, and residence at lower elevation as significant predictors. Notably, national *P. falciparum* prevalence decreased 53% relative to the parallel 2014-15 RDHS study, despite reports of increasing ART-R-associated mutations in Rwanda. These findings demonstrate the utility of ultra-sensitive molecular surveillance, and suggest that national malaria control efforts have substantially reduced malaria burden in Rwanda even amid rising antimalarial parasite prevalence. Subsequent studies on this data set will provide measures of minor *Plasmodium* species prevalence, as well as temporospatial analysis of antimalarial resistance markers in *P. falciparum* positive samples.

## Introduction

Malaria cases in the WHO Africa region have declined substantially since 2000 as a result of pointed efforts to reduce malaria morbidity and mortality[1]. Despite this progress, malaria remains endemic across much of the region, including in Rwanda, with *Plasmodium falciparum* (*P. falciparum*) being the most prevalent and deadly malaria-causing parasite species[1]. After major reductions in malaria burden between 2000 and 2012[2], Rwanda experienced a sharp resurgence between 2012 and 2018, with reported cases rising from approximately 640,000 in 2010 to 3.4 million in 2016[3]. In response, the country scaled up control interventions, reducing case numbers to 2 million in 2020[4] and 749,000 in 2023[5–7]. Malaria control remains challenging due to multiple emerging threats in Rwanda and the surrounding region such as increases in temperature and rainfall driven by climate change[8,9], increased pyrethroid insecticide resistance[10–12], and antimalarial resistant parasites[13–17]. Parasite antimalarial resistance may already be playing an outsized role[16,18,19]. In 2020, Rwanda was the first country in the WHO African Region to detect artemisinin partial resistance-mediating *PfKelch13* mutations[20,21] in samples from 2014[15] which threatens the progress made to reduce malaria burden. The sixth Rwandan Demographic and Health Survey (RDHS) in 2019-20 surveyed a nationally representative sample of respondents, collecting data intended to evaluate and improve public health within Rwanda[22]. Data was collected between November 9, 2019 and July 20, 2020, with a pause between March 21 and June 7, 2020 due to national lockdowns for the COVID-19 pandemic.

The overall focus of the RDHS 2019-20 was to provide updated data on demographic and health indicators to evaluate and assist policymakers on HIV, malaria, maternal health, and childhood nutrition programs[22]. Malaria surveillance efforts within the RDHS 2019-20 focused on women aged 15-49 years and children aged 6-59 months, with the RDHS Final Report showing a national malaria prevalence of <1% using both malaria rapid diagnostic tests (RDT) and microscopy to detect infection [22]. Previous research has demonstrated that molecular detection methods, such as quantitative real-time PCR (qPCR) can detect infections that were missed by conventional diagnostics, providing a more accurate estimate of malaria burden and epidemiology[23,24].

To better understand asymptomatic malaria during the emergence and rise of *Pfkelch13* mutations, we repurposed a representative subset of dried blood spot (DBS) samples from adult males and females originally collected in the 2019-20 RDHS for HIV testing, and utilized species-specific qPCR to detect *Plasmodium spp*. This work builds on a parallel study by Gaither et. al.[25] of the 2014-15 RDHS that used the same laboratory and statistical methodology, allowing for direct comparative analyses between these two DHS studies. Here, we focus on *P. falciparum* infection, assessing not only the change in distribution in Rwanda, but also the temporal changes in prevalence and demographic factors associated with infection in the 2014-15 and 2019-20 RDHS. Insights obtained from this study support the continuation of targeted interventions for malaria control in Rwanda, and act as the basis of subsequent studies that will evaluate *P. falciparum* positive samples for mutations in *Pfkelch13* and other parasite genes implicated in antimalarial drug resistance.

## Methods

### Ethics Statement

Residual dried blood spots (DBS) were provided after completion of the DHS by the Rwandan Ministry of Health and their use was granted by the Rwanda Biomedical Center (Reference 3543/RBC/2024). Use of DHS data for this project, including cluster geographic coordinates, was granted by the ICF and DHS Program. The IRBs at both the University of North Carolina and Brown University determined this study was within the realm of non-human subjects research.

### DHS Study Design

The 2019/20 RDHS surveyed 12,949 households within 500 GPS-located clusters-112 urban and 388 rural-defined as villages or part of villages identified by the 2012 Rwanda Population and Housing Census. Within each cluster, households were randomly assigned to either the Men’s Survey or Women’s Survey, with approximately 50% allocated to each. All women aged 15-49 and men aged 15-59 in the Men’s Survey were eligible for HIV testing, for which 13,942 DBS were collected. 50% of Men’s Survey households were selected for the first biomarker questionnaire, which included dual-method malaria testing-malaria RDT and blood smear microscopy-for women aged 15-49 and children under 5 years. Similarly, 50% of Women’s Survey households were selected for micronutrient testing and dual-method malaria testing for women and children in those households.

### Sampling Strategy

Our study utilized a nationally representative down-sampling strategy, which followed the previous 2014-2015 DHS analysis approach[25] as we aimed to provide both malaria prevalence data and sequence *P. falciparum* positive samples for mutations in genes implicated in antimalarial drug resistance. We randomly selected 50% of the residual DBS samples from each of 500 clusters for the random sample subset (n=6,963). We then identified clusters with high (>/=15%, n=16) or low (<15%, n=484) malaria prevalence using RDHS-reported RDT or microscopy-based *Plasmodium spp.* detection data (Fig 1). Samples from the 16 high-prevalence clusters that were not initially selected into the random sample subset (n=164) were added to the final data set. In total, 7,127 samples were tested for *P. falciparum* by qPCR and 5 samples were removed from analysis for indeterminate results.

**Fig 1.**
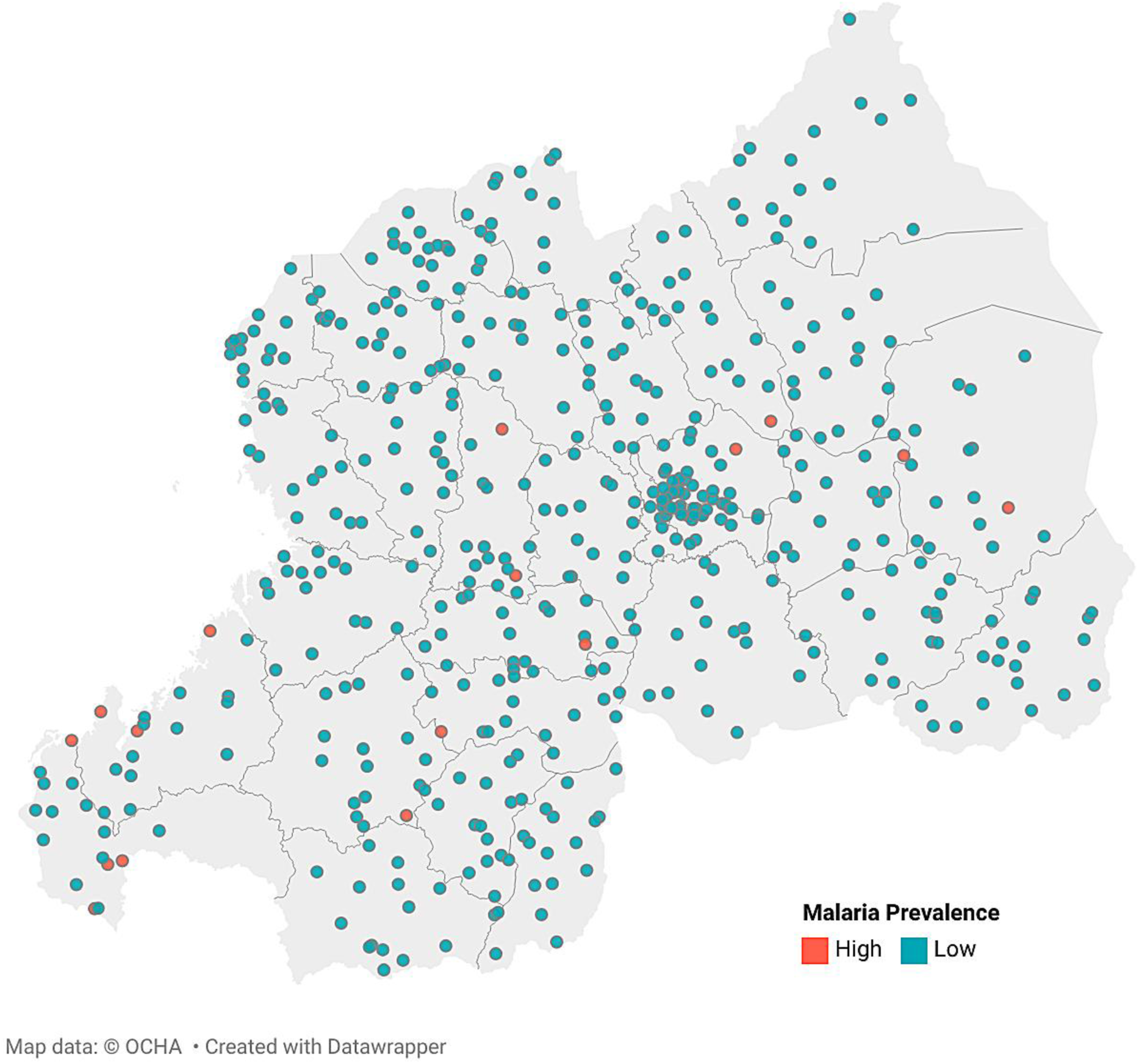
Distribution of 2019-20 Rwanda DHS cluster locations. Map of Rwanda depicting GPS-linked clusters in the 2019-20 RDHS with district boundaries included. Clusters are shaded based on *Plasmodium spp.* prevalence, high (≥15%, n=16) or low (<15%, n=484), as given by RDT or microscopy-based detection data from the RDHS 2019-20. Figure created with Datawrapper.com.

### Species-specific real-time quantitative PCR (qPCR)

We tested DNA from individual DBS samples for *P. falciparum* by real-time quantitative PCR (qPCR). DNA was extracted from three 6mm DBS punches using Chelex as previously described[26,27] and screened for *P. falciparum* infection using qPCR (BioRad) with primers specific to the 18S ribosomal gene of *P. falciparum* (dx.doi.org/10.17504/protocols.io.j8nlkyywwg5r/v1). Serial dilutions of the diagnostic plasmid containing the rRNA Gene (18s) from *P. falciparum* MRA-177 (BEI Resources, Manassas VA) were used as controls, with estimates for parasitemia based on an estimated six 18s rRNA gene copies per parasite metric[28,29]. All assays were run for 45 cycles to detect low-density infections. All data were analyzed using the BioRad Software (CFX Manager v2.1), with analysis of qPCR cycles 10-45, and utilization of the BioRad Software autocalculated baseline threshold and fluorescence drift correction. Positive samples were confirmed by manually reviewing amplification curves. Standard curves created by the serial dilutions of the control had a minimum R-squared value of 0.95 across all runs, and no false positives were detected in negative controls.

### Demographic and ecological variables

The DHS data is organized into recodes containing different variable sets. We utilized the household member, HIV, individual (women’s), and men’s recodes, merging the deidentified data from each recode based on DBS sample barcode and ID variables, as instructed in the Guide to DHS statistics DHS-8 (https://dhsprogram.com/data/Guide-to-DHS-Statistics/Analyzing_DHS_Data.htm). Then, qPCR data was linked to each individual based on the DBS sample barcode. The individual-level demographic variables assessed for association with malaria infection risk were sex, age, wealth quintile, education level, livestock ownership, source of drinking water, household bed net ownership, sleeping under an LLIN the night before the survey was performed, and whether the household met the World Health organization’s criteria for adequate bed net coverage of at least 1 net per 1.8 household members. Cluster-level variables included region, urban/rural status, elevation, month in which survey data was collected, average daily maximum temperature for the month in which survey data was collected, and precipitation levels during the month prior to survey data collection.

We utilized the geographic recode dataset, which contains geographic coordinates of each cluster. Cluster geographic coordinates are regarded as an estimated center of a cluster, provided within a 2km or 10km buffer zone for urban or rural clusters, respectively. Precipitation values were obtained from the Climate Hazards Group InfraRed Precipitation with Station (CHIRPS) dataset and cluster coordinate buffer zones were not included[30]. Cluster-level temperature values were obtained from the 2019-20 Rwanda DHS geospatial covariates recode which contains population, climate, and environmental variables.

### Statistical analyses

*P. falciparum* prevalence was estimated at national and district levels, with HIV sampling weights, inverse propensity of selection weights, and transmission intensity weights multiplied to produce a final weight for each sample. HIV sampling weights were provided by the RDHS and were based on sex and cluster, accounting for potential bias introduced due to the age and household selection constraints for eligibility for DBS collection for HIV testing. Inverse propensity for selection weights were created to correct for selection bias and equalize representation of the following important covariants: sex, age interval, education level, livestock ownership, bed net ownership, wealth index quintile, LLIN usage, month of data collection, cluster, region, urban/rural status, altitude, temperature during month of interview, and precipitation during month before interview. Transmission intensity weights were calculated based on the cluster status of high or low malaria prevalence, determined as previously described. This weighting strategy enables the use of covariate analysis to produce metrics that are representative of both the full DHS survey and national population. Data analysis was performed on R 4.4.1 (R Foundation for Statistical Computing) using the rdhs (0.8.4), svyr (1.3.0), sf (1.0-16), and terra (1.8-54), rnaturalearth (1.0.1), ggplot2 (3.5.1) packages. All analysis code is available on GitHub (github.com/jzuromski/RW2019DHS).

## Results

### Study Population

Our molecular testing study sample set (n=7,127, weighted n= 7,122) was 53% female and 38% aged 15-24 years, with 80% living in rural areas. 68% had a primary school education or no education, compared to 75% in the 2014-15 RDHS[25]. The rate of household bed net ownership was 69%, with 50% of the study population living in a household that meets the WHO criteria of at least 1 net per 1.8 household members, and 59% of respondents reporting sleeping under a long-lasting insecticide-treated net the night before the survey. Comparison of our molecular testing study sample set (n= 7,127, weighted n= 7,122) to all RDHS DBS samples (n=13,941, weighted n=14,110) displayed representativeness across our key characteristics of interest, including sex, age, residence, and wealth index (Table 1).

**Table 1:**
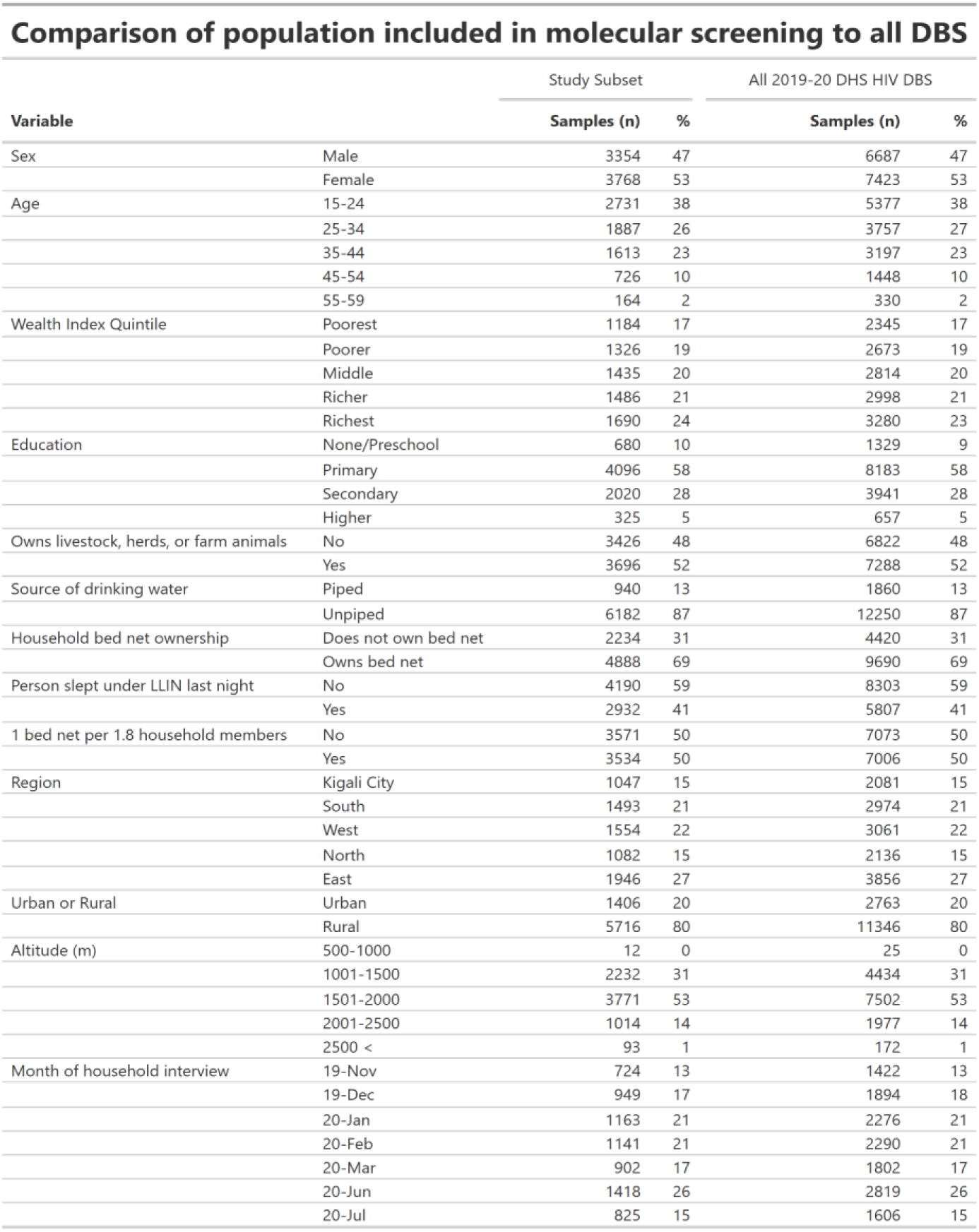
Comparison of population included in molecular screening to all DBS. Weighted sample counts and percentage of individuals with a demographic characteristic compared to the total within the variable. Measures show that the population included in molecular screening is representative of all individuals who provided DBS for HIV testing in the 2019-20 DHS.

### Pf prevalence by qPCR

A total of 616 *P. falciparum* infections were detected in 7,127 samples (unweighted). The weighted national *P. falciparum* prevalence was 7.7% (95%CI, [6.79%, 8.70%]) using a qPCR cycle threshold (CT) of 45 and applying sampling weights, as previously described. District-level weighted Pf prevalence ranged from 0.0% in Musanze to 17.4% in Ruhango (Fig 2a).

**Fig 2:**
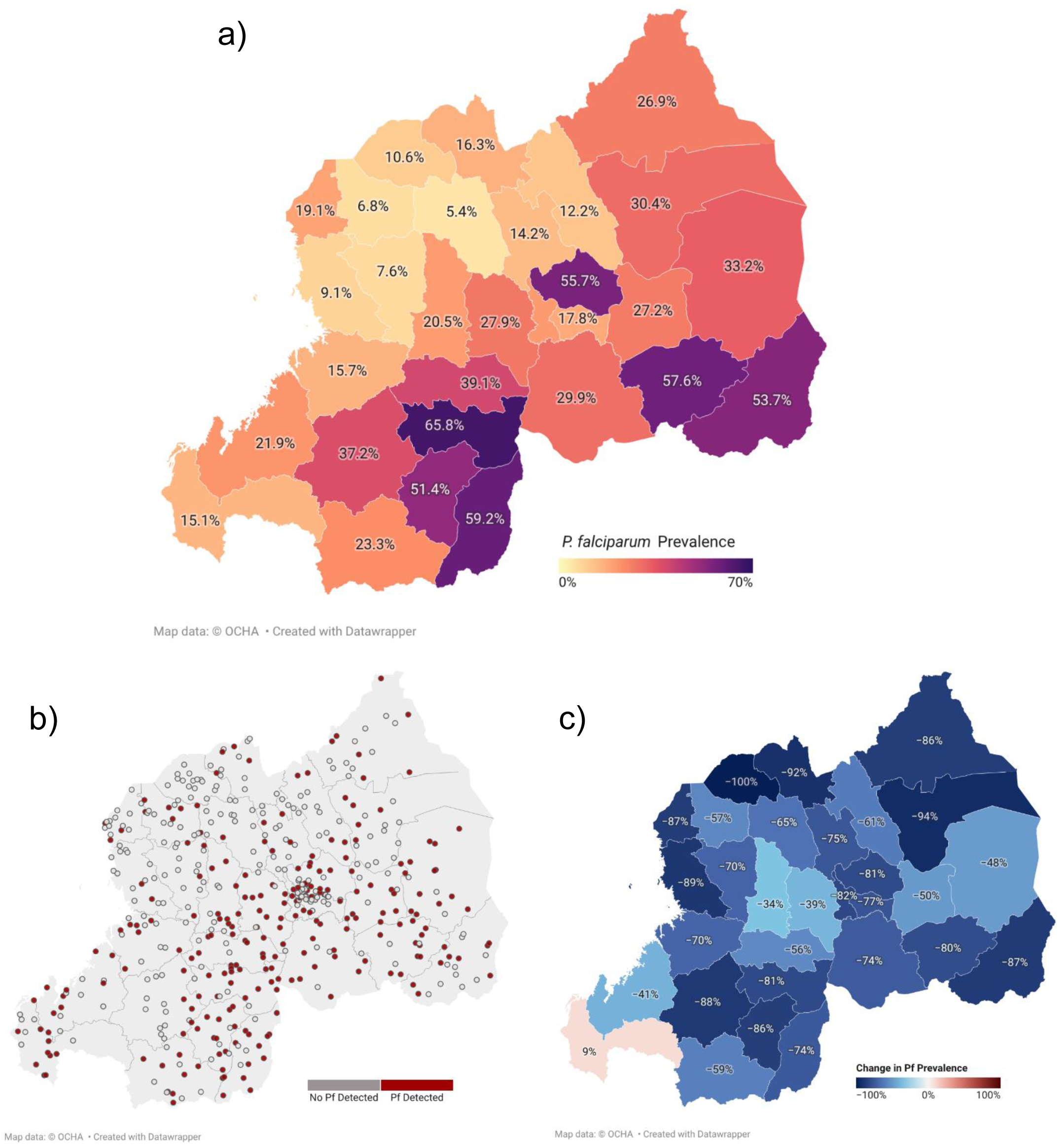
*P. falciparum* prevalence. Detection of *P. falciparum* within our representative sample set was accomplished using Pf-specific 45-cycle qPCR assays on DNA extracted from dried blood spots (DBS) obtained from individuals surveyed in the RDHS 2019-2020. **a)** District-level weighted *P. falciparum* prevalence in Rwanda 2019-20 DHS. Data are weighted as previously described and provided as the percentage of samples positive for Pf using a qPCR cutoff of 45 cycles. **b)** Distribution of clusters where Pf was detected. GPS-provided locations of all 500 clusters are provided, with those shaded red indicating *P. falciparum* detected in one of more samples and clusters outlined in grey having no samples with detected *P. falciparum*. **c)** Temporospatial analysis of change in *P. falciparum* prevalence between 2014/15 and 2019/20 RDHS. District-level change in weighted *P. falciparum* prevalence between the 2014-15 Gaither et. al[25] and present 2019-20 RDHS studies. Districts are shaded according to the magnitude and direction of change. Figure created with Datawrapper.com.

Within *P. falciparum* positive samples, the median parasitemia was 7.3 parasites/uL, with the vast majority (77%) of positive samples having under 100 parasites/uL (Fig 3). Median parasitemia was 7.3 parasites/uL, with an interquartile range of 1.13 - 77.54 parasites/uL.

**Fig 3:**
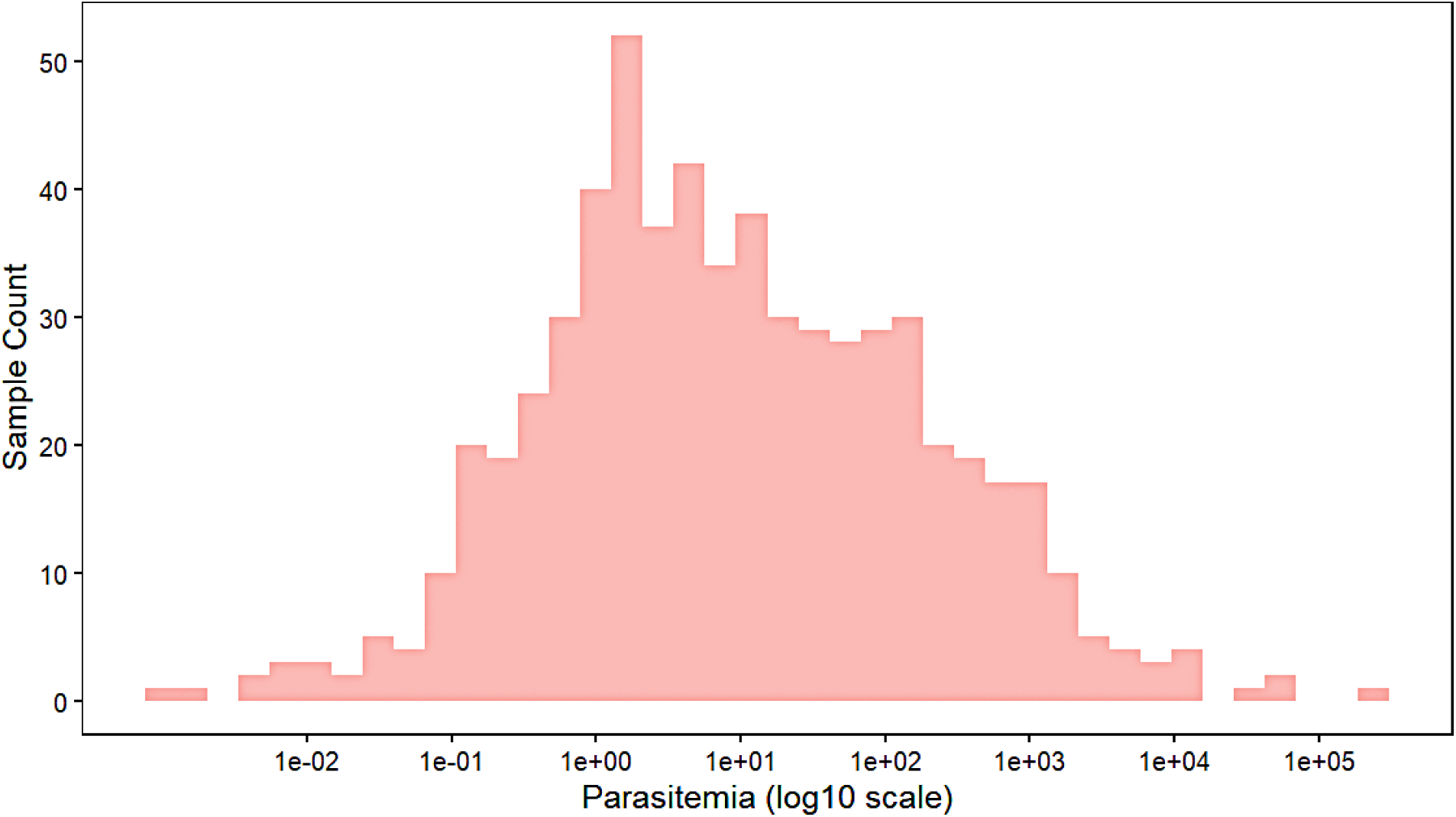
Parasitemia estimates for *P. falciparum* positive samples. Starting parasitemia (parasites/uL) was calculated based on our standard curve derived from serially diluted *P. falciparum* rRNA 18S diagnostic plasmid, as previously described.

A CT cutoff of ≤40 was used to restrict positive counts to samples with approximately 1 parasite or greater per microliter of blood, providing a more conservative national weighted *P. falciparum* prevalence estimate of 7.3% (95%CI [6.4%, 8.2%]). At a CT of 45, a total number of 616 (unweighted, weighted n=547) *P. falciparum* infections were detected, compared to 585 (unweighted, weighted n=517) at a CT cutoff of ≤40, reflecting the detection of very few ultra-low-density parasite infections. When using the CT cutoff of ≤40, district-level weighted *P. falciparum* prevalence decreased by a mean of 0.4%, median of 0.39%, and a maximum of 1.7% (Table 2).

**Table 2:** District-level *P. falciparum* prevalence by qPCR cycle cutoff. Weighted sample counts and district-level *P. falciparum* prevalence were calculated using qPCR cycle cutoffs of 45 and ≤40. Relative differences in prevalence measures were calculated as the difference between prevalence at 45 and ≤40 and are shaded by difference in prevalence measures when samples with CT between 40 and 45 are removed. Total represents the sum (n) or mean (%) of rows.

| District-level Pf prevalence by qPCR cycle cutoff |  |  |  |  |  |  |
| --- | --- | --- | --- | --- | --- | --- |
| District | Samples (n) | 45 cycles |  | 40 cycles |  | Δ Pf Prevalence |
|  |  | Pf Count (n) | Pf Prev (%) | Pf Count (n) | Pf Prev (%) |  |
| Bugesera | 272 | 21 | 7.87 | 19 | 6.94 | 0.93 |
| Burera | 207 | 3 | 1.38 | 3 | 1.29 | 0.09 |
| Gakenke | 190 | 4 | 1.90 | 4 | 1.90 | 0.00 |
| Gasabo | 531 | 58 | 10.86 | 56 | 10.56 | 0.30 |
| Gatsibo | 319 | 6 | 1.81 | 5 | 1.55 | 0.26 |
| Gicumbi | 241 | 12 | 4.79 | 12 | 4.79 | 0.00 |
| Gisagara | 182 | 28 | 15.14 | 27 | 14.60 | 0.54 |
| Huye | 170 | 13 | 7.42 | 12 | 6.86 | 0.56 |
| Kamonyi | 232 | 39 | 16.97 | 38 | 16.51 | 0.46 |
| Karongi | 194 | 9 | 4.76 | 8 | 4.29 | 0.47 |
| Kayonza | 263 | 45 | 17.12 | 41 | 15.69 | 1.43 |
| Kicukiro | 301 | 13 | 4.17 | 13 | 4.17 | 0.00 |
| Kirehe | 232 | 16 | 6.85 | 15 | 6.29 | 0.56 |
| Muhanga | 196 | 27 | 13.59 | 26 | 13.13 | 0.46 |
| Musanze | 269 | 0 | 0.00 | 0 | 0.00 | 0.00 |
| Ngoma | 278 | 33 | 11.73 | 28 | 10.18 | 1.55 |
| Ngororero | 203 | 5 | 2.26 | 4 | 1.73 | 0.54 |
| Nyabihu | 209 | 6 | 2.95 | 5 | 2.31 | 0.64 |
| Nyagatare | 338 | 13 | 3.78 | 11 | 3.14 | 0.64 |
| Nyamagabe | 179 | 8 | 4.41 | 8 | 4.41 | 0.00 |
| Nyamasheke | 220 | 28 | 12.96 | 28 | 12.68 | 0.29 |
| Nyanza | 193 | 24 | 12.50 | 24 | 12.19 | 0.30 |
| Nyarugenge | 214 | 8 | 3.81 | 8 | 3.81 | 0.00 |
| Nyaruguru | 151 | 14 | 9.50 | 12 | 7.76 | 1.75 |
| Rubavu | 306 | 8 | 2.56 | 8 | 2.56 | 0.00 |
| Ruhango | 189 | 33 | 17.36 | 33 | 17.36 | 0.00 |
| Rulindo | 176 | 6 | 3.51 | 6 | 3.51 | 0.00 |
| Rusizi | 206 | 34 | 16.44 | 33 | 15.74 | 0.70 |
| Rutsiro | 216 | 2 | 0.99 | 2 | 0.99 | 0.00 |
| Rwamagana | 245 | 34 | 13.69 | 32 | 13.01 | 0.69 |
| **Total** | **7,122** | **547** | **7.77** | **517** | **7.33** | **0.44** |

*P. falciparum* was detected in 49% (244 of 500) clusters, with only 23% of clusters in the Northern Province and 37% of clusters in the Western Province having detectable malaria (Fig 2b). *P. falciparum* was detected in 46% of clusters in Kigali City and in 60% and 68% of clusters in the Eastern and Southern provinces, respectively.

### Temporal trends: 2014-15 vs 2019-20

Analyses between the results from the 2014-15 RDHS qPCR study by Gaither et. al.[25] and the present study were possible due to the use of comparable subsampling schemes, laboratory testing protocols, and data analysis approaches. There is a clear decline in national *P. falciparum* prevalence from 16.3% (95%CI [14.5%, 18.1%]) in the 2014-15 RDHS to 7.7% (95%CI [6.8%, 8.7%]) in the 2019-20 RDHS. Between 2014-15 and 2019-20 RDHS, district-level weighted *P. falciparum* prevalence decreased in all districts except Rusizi (8.85% increase) (Fig 2c). Only 11 districts had a Pf prevalence greater than 10% in the 2019-20 RDHS, compared to 26 districts in the 2014-15 RDHS (Gaither et. al., Supplemental Table 6[25]).

### Determinants of Pf Infection

Examining the weighted prevalence of *P. falciparum* for a number of variables without adjusting or controlling for other factors, *P. falciparum* prevalence was higher in males at 9.5% (95%CI, [8.24%, 10.96%]) compared to 6.1% in females (95%CI, [5.16%, 7.10%]), (Table 3). *P. falciparum* infection was negatively associated with variables that indicate higher socioeconomic status, such as higher wealth, increased educational attainment, bed net ownership, and piped drinking water (Table 3). Cluster-level factors with higher *P. falciparum* infection rates included residence in the Southern Province, rural areas, at an altitude between 1001 and 1500m, and in clusters with monthly average temperatures at or above the average temperature of all clusters.

**Table 3:**
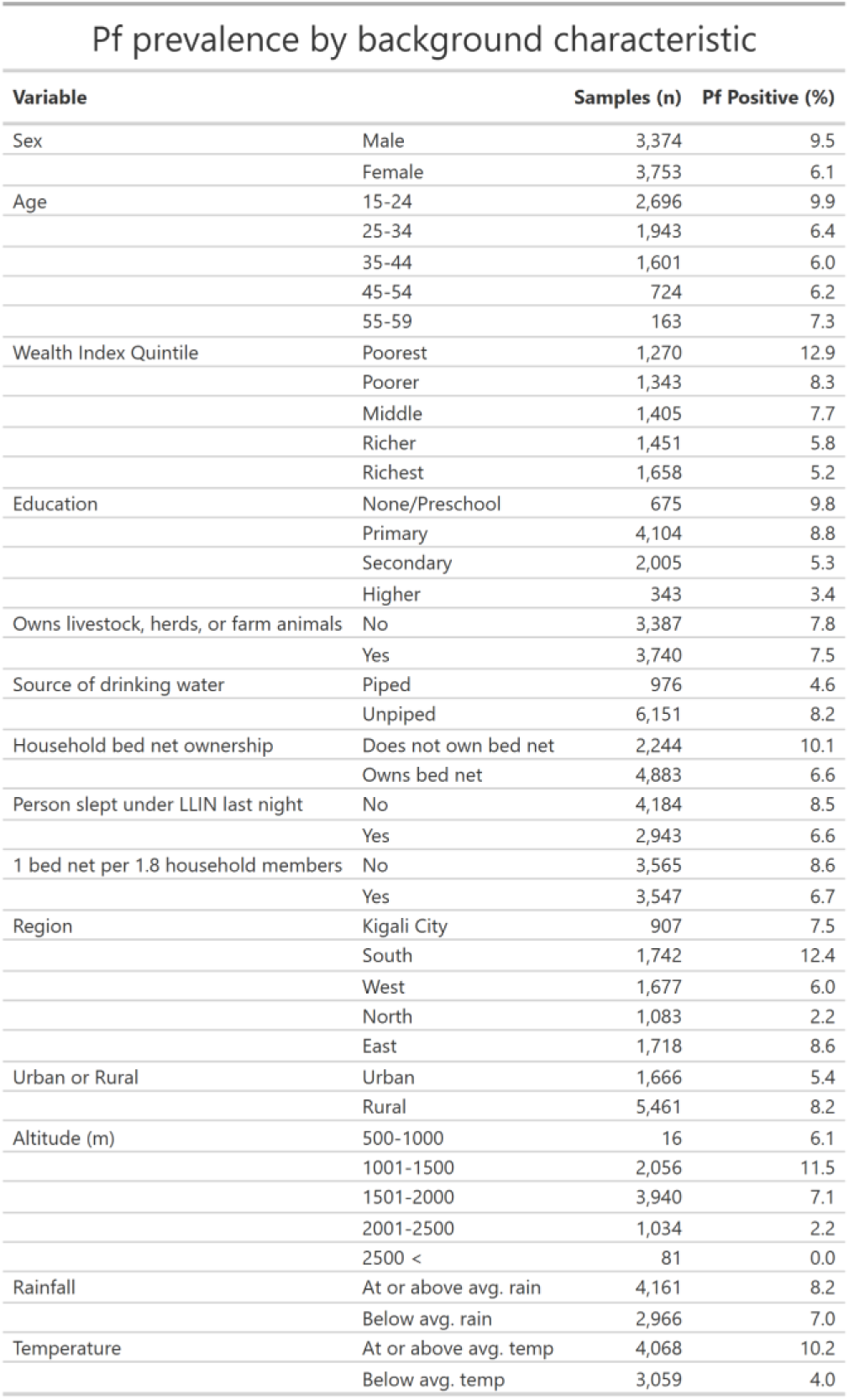
Pf positivity rates by study population characteristic. For each variable, unweighted sample counts are reported with *P. falciparum* positivity rates, which are weighted as previously described.

Several risk factors for qPCR-detected *P. falciparum* infection were detected using bivariate association analyses with survey data weighted as described previously. Variables with greater than two categories or levels were transformed into binaries to compare using a linear model. Lower prevalence of *P. falciparum* malaria was found in females, individuals with secondary or higher educational attainment, bed net owners, and those residing in a cluster that was located at an elevation above 1500m or had below average monthly temperatures (Fig 4). A one-year change in age did not prove to be a risk factor for *P. falciparum* infection, whereas individuals in the 14-24 age group were significantly more likely to be infected compared to those over 24 years old. Livestock ownership and the cluster’s prior month average rainfall did not show significant association with *P. falciparum* infection.

**Fig 4:**
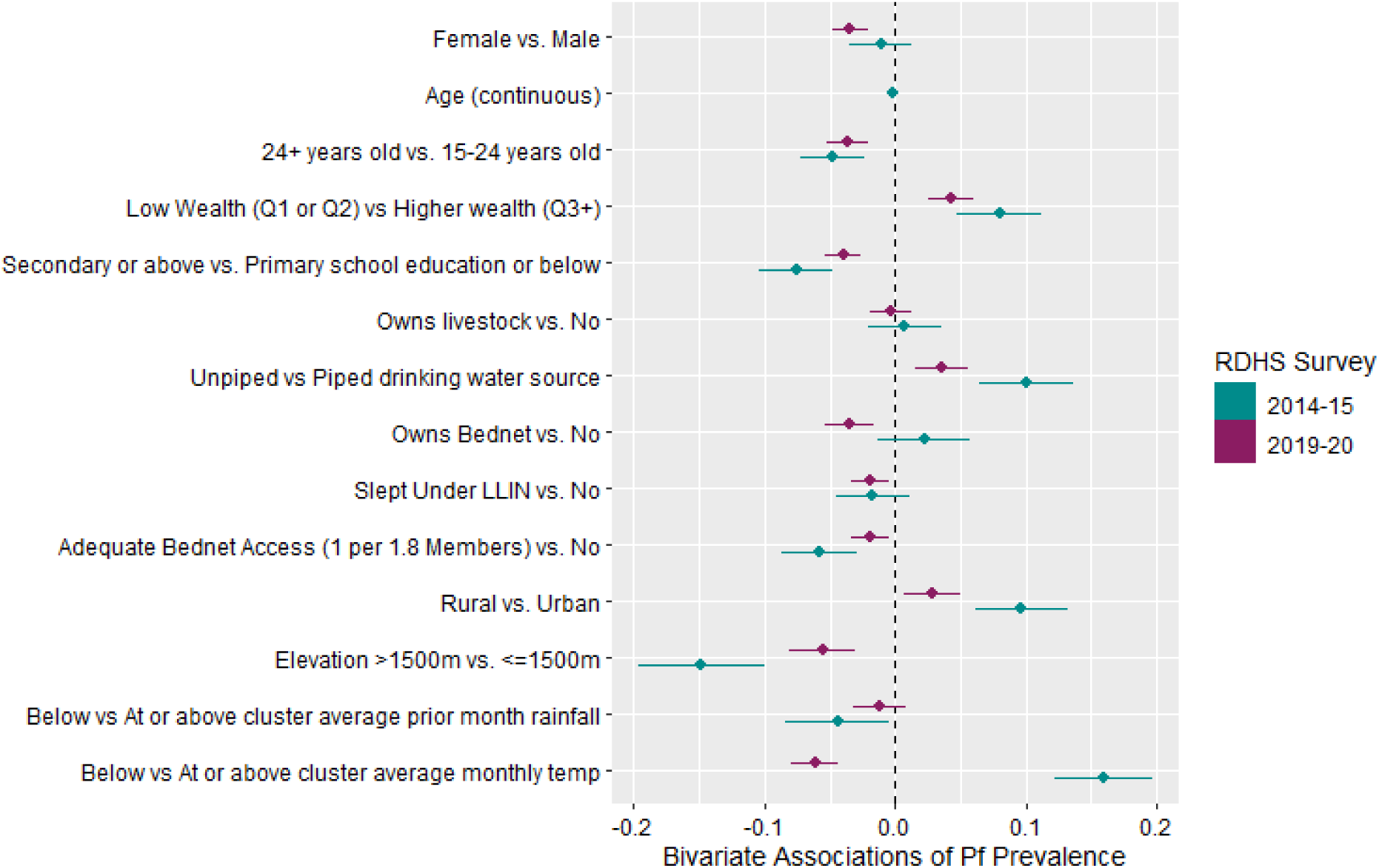
Bivariate associations between *P. falciparum* prevalence and demographic or environmental factors in the 2014-15 and 2019-20 RDHS. Forest plot showing weighted bivariate associations between *P. falciparum* prevalence (measured by qPCR) and key demographic and environmental variables. Results from the 2014-15 RDHS (green) are reproduced directly from Gaither et al. Fig 4[25], using their original point estimates and confidence intervals. Results from the current 2019-20 RDHS analysis (purple) were generated using the same statistical approach to enable direct head-to-head comparison. For each variable, the plot displays the estimated difference in *P. falciparum* prevalence relative to the reference category (the second category listed), along with corresponding 95% confidence intervals.

## Discussion

Malaria control measures in Rwanda have expanded considerably in recent years, with an increase in investment in malaria control measures such as ITN and IRS campaigns, no-cost malaria detection, and treatment services through community health workers[5,7]. We assessed the rate and risk factors of *P. falciparum* infection in Rwanda using a nationally representative RDHS 2019-20 sampler set that included both dried blood spot (DBS) samples and demographic data from asymptomatic Rwandan adults. This study directly parallels the molecular survey by Gaither et al. of the 2014-15 RDHS, which applied the same laboratory and statistical methodologies, thus permitting direct comparison between the two national surveys.

We found a national *P. falciparum* prevalence of 7.7% in 2019-20, a substantial reduction from 16.3% prevalence found by Gaither et. al.[25] in the 2014-15 RDHS. The Eastern and Southern provinces had the highest malaria burden, consistent with historical trends[31,32]. District-level prevalence comparisons showed significant reductions in all districts except Rusizi (Fig 2c), which is regarded as a high-burden district[31]. While a nearly 50% national decline reflects the success of Rwanda’s investment in malaria control, the detection of *P. falciparum* within all districts except Musanze highlights the need for strategies to address malaria transmission. Our findings that sex, wealth level, educational attainment, bed net ownership and use, elevation, and high monthly temperature were predictors of *P. falciparum* infection, which is consistent with findings from the 2014-15 RDHS[27], as well as others showing environment[9,9,33] and socioeconomic status[34,35] are predictors of malaria infection. Age as a continuous measure did not show any association with *P. falciparum* infection, but when dichotomized, being aged 15-24 was significantly associated with infection versus being over 24 years of age.

We combined the bivariate regression model used by Gaither et. al. with our data to produce a direct comparison of individual associations based on prevalence measures in the 2014-15 and 2019-20 RDHS. Interestingly, our bivariate analyses of key demographic and environmental risk factors for 2019-20 RDHS *P. falciparum* prevalence were less predictive compared to the same analysis rendered on the 2014-15 RDHS (Fig 3). This reflects success by the 2016-2018 Malaria Operational Plans, a multi-level intervention strategy that included mass ITN distribution for universal coverage in 2016/17, IRS campaigns in 11 high burden districts, community-based treatment, and free malaria diagnosis and treatment for economically vulnerable populations[31]. These pointed decisions by the Rwanda National Malaria Control Program and its partners likely reduced the effect of these risk factors compared to the 2014-15 RDHS[25]. In addition, monthly temperatures and bed net ownership had an inverse effect between the 2014-15[25] and 2019-20 RDHS.

Consistent with findings from the earlier RDHS molecular study, prevalence measured by qPCR was substantially higher than that detected using RDT and microscopy by the RDHS program. In the 2019–20 RDHS, *P. falciparum* prevalence among women aged 15–49 years was 0.5% and 1.2% by microscopy and RDT, respectively, compared to qPCR positivity rate of 6.1% (95%CI [5.2%, 7.1%]) from samples from females in the same age group. Therefore, while microscopy and RDTs are invaluable for symptomatic cases, they are insufficient for detecting the full burden of infection at the population level. This discrepancy illustrates the limitations of routine surveillance tools for detecting the true burden of asymptomatic and low-density infections, particularly during low-transmission seasons when RDHS samples are collected.

qPCR is a highly sensitive approach, enabling the detection of low-density infections by running assays for 45 cycles. We ran analyses with a cycle threshold (CT) of 40 cycles, which limits positivity to samples with approximately 1 parasite or greater per microlitre of blood. *P. falciparum* prevalence using a CT ≤40 cutoff was 7.3% (95%CI [6.4%, 8.2%]), which is comparable to the 7.7% prevalence in data collected for up to 45 qPCR cycles. At the district level, the difference in prevalence when utilizing a CT ≤40 cut off was limited. This reflects the detection of very few ultra-low-density parasite infections and provides evidence to support our confidence in the 45-cycle data.

Finally, we note several limitations. Cluster sample sizes were insufficient for precise cluster-level estimates, with a minimum of 7, maximum of 34, and median of 14 samples tested for *P. falciparum* in each cluster. The low *Plasmodium spp.* prevalence (1% nationally) detected by the RDHS made determination of high and low prevalence clusters more difficult. One could argue that the threshold for high prevalence could, therefore, have been shifted to 10% for this study rather than maintaining it at 15% to align with the parallel 2014/15 RDHS study[25]. Pf-specific qPCR was utilized within this analysis, and the qPCR positivity threshold of 45 cycles, while extremely sensitive, may detect non-transmissible infections. Nonetheless, the patterns observed are consistent, biologically plausible, and highly informative to ongoing malaria elimination efforts.

The use of qPCR enabled the detection of low-density infections in samples collected outside of peak malaria transmission season, which is critical to understanding the burden of *P. falciparum* during low or baseline malaria transmission seasons. Our studies underscore the limitations of routine surveillance tools and advocate for integration of molecular diagnostics and inclusion of adult males in future RDHS or similar surveys.

## Conclusions

Molecular surveillance using DHS-linked qPCR highlights a hidden reservoir of *P. falciparum* infection within adults across Rwanda. Despite progress in case reduction, substantial submicroscopic infections remain, particularly within the southern half of the country. Our results strongly support the incorporation of molecular diagnostics into malaria surveillance systems and national health surveys to better guide elimination strategies.

## Data Availability

All data produced in the present work are contained in the manuscript. R analysis scripts can be found at github.com/jzuromski/RW2019DHS. Sample sets can be acquired with permission from the DHS.

## Acknowledgements

We would like to thank the Ministry of Health-Rwanda for providing access and approval of DBS for this study. The following reagent was obtained through BEI Resources, NIAID, NIH: Diagnostic Plasmid Containing the Small Subunit Ribosomal RNA Gene (18S) from *Plasmodium falciparum*, MRA-177, contributed by Peter A. Zimmerman.

Funding: K24AI134990 to JJJ, R01 AI 156267 to JAB

